# Analysis of Risk Factors and the Establishment of a Predictive Model for Thrombosis in Patients with immune thrombocytopenia

**DOI:** 10.1101/2024.08.21.24312388

**Authors:** Hui Liang, Lingxue Duan, Manyu Long, Songyuan Tie, Changyan Sun, Sha Ma, Jing Wang, Shuya Wang

## Abstract

**Objectives:** To explore the risk factors for thrombi occurring in patients with immune thrombocytopenia (ITP) and establish a risk prediction model to better predict the risk of thrombosis in patients with ITP.

**Methods:** We retrospectively analyzed 350 ITP patients who had been hospitalized in The First People’s Hospital of Yunnan Province between February to June 2024. For all patients, we recorded demographic characteristics and clinical data, analyzed the risk factors for thrombosis in ITP patients and then developed a risk prediction model.

**Results:** Stepwise logistic regression analysis indicated that a high complement D-dimer level, a low PLT and a high Padua score were independent risk factors for thrombosis in ITP patients. According to multivariate analysis, a predictive model for thrombus risk was successfully established; the area under the ROC curve(AUC) was 0.673 (95% CI: 0.615-0.730) and the maximum Youden index, sensitivity and specificity were 0.272, 47.0% and 80.2%, respectively.

**Conclusion:** A high complement D-dimer level, low PLT level, and a high Padua score were shown to be independent risk factors for thrombosis in ITP patients. We developed a risk prediction model based on these three risk factors that could predict the risk of thrombosis in ITP patients to some extent.

## Introduction

Immune thrombocytopenia (ITP) is an autoimmune disease characterized by peripheral platelet destruction and insufficient platelet production[1], at the same time, predisposing patients to an increased risk of bleeding[2]. The current research generally believes that its pathogenesis involves not only cellular immune disorders and humoral immunity of the disorder, that also involves aspects such as oxidative stress in the body[3]. But these studies that ITP itself may be a potential thrombogenic risk disease.The incidence of thrombus is higher than that of normal population, even though the platelet level is low that the thrombotic events also may be occurred[4]. Incongruously, patients with ITP may be at risk of thrombosis; large-scale population-based studies found increased rates of both venous thromboembolism (VTE) and arterial (ATE) thromboembolism in patients with ITP compared with age and gender-matched individuals without ITP[5, 6]. We found that systematic reviews and meta-analyses that provided supportive evidence that ITP may have a prothrombotic tendency influenced by therapy-specific factors, in particular thrombopoietin receptor agonists (TPO-RAs)[7], but new research suggests otherwise[8, 9]. To date, delineating the risk factors for thrombosis in ITP remains incomplete but likely encompasses cumulative individual, ITP disease-specific, and ITP therapy-related risk factors[10, 11]. At present, Thrombotic risk has not been evaluated in detail in Chinese ITP patients. Therefore, the main purpose of this study was to investigate thrombotic risks for Chinese ITP patients. Our team retrospectively collected clinical data of 350 patients with ITP hospitalized. The thrombogenesis and influencing factors of ITP are intended to enrich the information about ITP thrombogenesis, the relevant data of health conditions can provide help for the formulation of clinical prevention strategies.

## Methodology

### Date of access to data

Data accessed on June 2024 and authors had access to information that could identify individual participants during or after data collection.

### Patients

All patients diagnosed ITP who were added to the ITP repository at the First People’s Hospital of Yunnan Province (February to June, 2024) were included in the study. Patients without detailed documentation of their diagnosis and each subsequent relapse, alongside data on their demographic features, comorbidities, medications, family history, and baseline organ function, were excluded.

All patients met the Consensus of Chinese experts on the diagnosis and treatment of adult primary immune thrombocytopenia (version 2016). Baseline clinic data, including the demographics, such as age, gender, C-creative protein(CRP), erythrocyte sedimentation rate(ESR), clinical manifestation, and laboratory results of each patient at first diagnosis were collected, including inflammatory markers, routine blood results, renal function, blood fat, complement C3, complement C4,; the Padua rating scale to evaluate the risk of thrombi[12], lupus anticoagulant (LA), Anti-cardiolipin antibody-IgM/IgG (aCLIgM/IgG), Anti-β 2-glycoprotei Ⅰ antibody (aβ2-GPⅠ) and so on.

### Ethics statement

This study was conducted following the Declaration of Helsinki and approved by the Ethics Committee of the First People’s Hospital of Yunnan Province (KHLL2024-KY171), it did not involve animal or human clinical trials. And data collection was based on complete medical records and data analysis was performed anonymously. All our research methods were in accordance with relevant guidelines and regulations.

### Statistical analysis

The data were analyzed using SPSS version 26.0 (SPSS Inc., Chicago, IL, USA). Patient characteristics and factors were analyzed using t-test and chi-square tests. Measurement data that do not conform to the normal distribution are expressed as M (P25, P75).The rank sum test was used to compare the two groups. performed uni-variate analysis to examine the difference between the thromboembolism group and the non thromboembolism group. Apply binary logistic regression modeling technique to analyze risk factors for thromboembolism ITP patients. Variables that had a p-value of < 0.05 in uni-variate analysis were selected into the multi-variable logistic analysis to get further independent risk factors. In the multi-variable logistic analysis, variables with a P value of < 0.05 were identified to be the independent risk factors automatically and finally selected into the final model. Regression coefficients were used to generate prognostic nomograms. Model discrimination was measured quantitatively with the concordance index. The risk prediction model was subsequently established pursuant to the risk factors determined by multi-factor logistic regression analysis. Receiver operator characteristic curve (ROC) analysis and DeLong inspection were used to evaluate the predictive value of the risk prediction model. P < 0.05 was deemed to be of statistical significance.

## Results

### Patient characteristics

We analyzed a total of 350 patients diagnosed with ITP, the patients from the First People’s Hospital of Yunnan Province(included Rheumatology and immunology, Hematology department and General surgery department) during the time period from February to June, 2024. Patients with repeated hospitalization are excluded according to the patient’s admission certificate.The median (IQR) age at ITP diagnosis was 45 (30-57) years, Only 14 cases were male and the rest were female. The baseline characteristics and main clinical features of the study are shown in Table 1. Primary ITP comprised the majority of ITP diagnoses are 96 cases, while autoimmune disorders were the most common secondary ITP. Of the study, the 315 cases of combined with Autoimmune disorder, including connective tissue disease 102 cases, systemic lupus erythematosus 64 cases, anticardiolipin syndrome in 41 cases, and Sjogren’s syndrome in 108 cases. In terms of drug therapy, hormone therapy was the main treatment, accounting for 90.29%, meanwhile included immunosuppressants (56.29%), intravenous immunoglobulin (25.43%), platelet transfusion (28%), TPO therapy (26.57%) and so on.

**Table 1.** Comparison of General Information and Clinical Manifestation.

| Characteristics | TE (n=116) | No TE (n=234) | Z/X<sup>2</sup> | P |
| --- | --- | --- | --- | --- |
| Sex, male/female, n | 13/103 | 1/233 | 23.468 | <0.001 |
| Age(y), median (IQR) | 38.50 (26-54.75) | 47 (33-58) | -2.534 | 0.11 |
| Ethnicity, n |  |  |  |  |
| Han nationality | 108 | 203 | 3.160 | 0.075 |
| Non-Han nationality | 8 | 31 |  |  |
| Height(cm<sup>2</sup> ), median (IQR) | 158 (156-163) | 158 (155-160) | -1.442 | 0.149 |
| Weight(kg ), median (IQR) | 56 (52-65.75) | 54 (49-60.125) | -2.814 | 0.005 |
| Course of a disease(M), median (IQR) | 6.5 (1.00-45.00) | 12.0 (1.00-60.00) | -0.862 | 0.388 |
| Primary/Secondary ITP, n | 26/90 | 70/156 | 2.782 | 0.095 |
| Combined with Autoimmune disorder |  |  |  |  |
| CTD | 47/27 | 109/75 | 0.403 | 0.525 |
| SLE | 58/16 | 136/48 | 0.564 | 0.453 |
| APS | 56/18 | 161/23 | 5.520 | 0.019 |
| SS | 40/34 | 110/74 | 0.712 | 0.399 |
| Medication |  |  |  |  |
| Corticosteroids | 14/102 | 20/214 | 1.097 | 0.295 |
| TPO-RAs | 82/34 | 175/59 | 0.667 | 0.414 |
| immunosuppressive agents | 67/49 | 86/148 | 13.909 | <0.001 |
| intravenous immunoglobulin | 80/36 | 181/53 | 2.875 | 0.090 |
| platelet infusion | 76/40 | 176/58 | 3.617 | 0.057 |
| anticoagulant | 110/6 | 227/7 | 1.031 | 0.310 |

Abbreviations: TE, thromboembolism; IQR, interquartile range; CTD: Connective tissue disease; SLE: Systemic lupus erythematosus; APS: antiphospholipid syndrome; SS: Sjogren syndrome; TPO: thrombopoietin.

In terms of laboratory results, Erythrocyte sedimentation rate (ESR), Fibrinogen(FIB) and degradation products(FDP) were significantly higher in the ITP thrombus group than those in the ITP group without thrombi (P < 0.001, P = 0.005 and P = 0.027, respectively). The levels of Platelet, neutrophil count and Padua scores swere significantly lower than those in the ITP group without thrombi (P = 0.028, P< 0.001, and P<0.001, respectively)(Table 2).

**Table 2.**
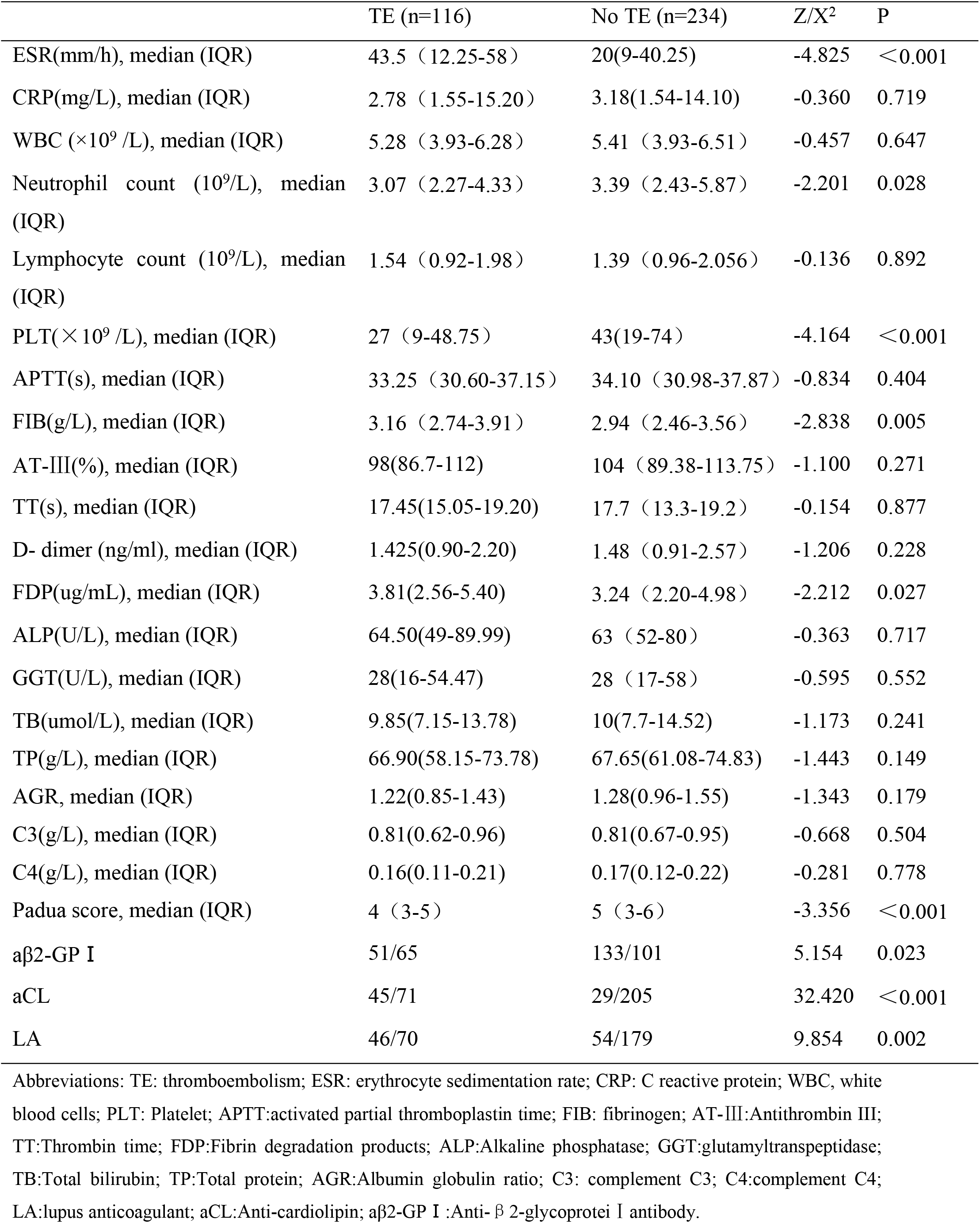
Comparison of Laboratory Results and Padua Scores.

|  | TE (n=116) | No TE (n=234) | Z/X<sup>2</sup> | P |
| --- | --- | --- | --- | --- |
| ESR(mm/h), median (IQR) | 43.5 (12.25-58) | 20(9-40.25) | -4.825 | <0.001 |
| CRP(mg/L), median (IQR) | 2.78 (1.55-15.20) | 3.18(1.54-14.10) | -0.360 | 0.719 |
| WBC (×10<sup>9</sup> /L), median (IQR) | 5.28 (3.93-6.28) | 5.41 (3.93-6.51) | -0.457 | 0.647 |
| Neutrophil count (10<sup>9</sup>/L), median (IQR) | 3.07 (2.27-4.33) | 3.39 (2.43-5.87) | -2.201 | 0.028 |
| Lymphocyte count (10<sup>9</sup>/L), median (IQR) | 1.54 (0.92-1.98) | 1.39 (0.96-2.056) | -0.136 | 0.892 |
| PLT(×10<sup>9</sup> /L), median (IQR) | 27 (9-48.75) | 43(19-74) | -4.164 | <0.001 |
| APTT(s), median (IQR) | 33.25 (30.60-37.15) | 34.10 (30.98-37.87) | -0.834 | 0.404 |
| FIB(g/L), median (IQR) | 3.16 (2.74-3.91) | 2.94 (2.46-3.56) | -2.838 | 0.005 |
| AT-III(%), median (IQR) | 98(86.7-112) | 104 (89.38-113.75) | -1.100 | 0.271 |
| TT(s), median (IQR) | 17.45(15.05-19.20) | 17.7 (13.3-19.2) | -0.154 | 0.877 |
| D- dimer (ng/ml), median (IQR) | 1.425(0.90-2.20) | 1.48 (0.91-2.57) | -1.206 | 0.228 |
| FDP(ug/mL), median (IQR) | 3.81(2.56-5.40) | 3.24 (2.20-4.98) | -2.212 | 0.027 |
| ALP(U/L), median (IQR) | 64.50(49-89.99) | 63 (52-80) | -0.363 | 0.717 |
| GGT(U/L), median (IQR) | 28(16-54.47) | 28 (17-58) | -0.595 | 0.552 |
| TB(umol/L), median (IQR) | 9.85(7.15-13.78) | 10(7.7-14.52) | -1.173 | 0.241 |
| TP(g/L), median (IQR) | 66.90(58.15-73.78) | 67.65(61.08-74.83) | -1.443 | 0.149 |
| AGR, median (IQR) | 1.22(0.85-1.43) | 1.28(0.96-1.55) | -1.343 | 0.179 |
| C3(g/L), median (IQR) | 0.81(0.62-0.96) | 0.81(0.67-0.95) | -0.668 | 0.504 |
| C4(g/L), median (IQR) | 0.16(0.11-0.21) | 0.17(0.12-0.22) | -0.281 | 0.778 |
| Padua score, median (IQR) | 4 (3-5) | 5 (3-6) | -3.356 | <0.001 |
| aβ2-GP I | 51/65 | 133/101 | 5.154 | 0.023 |
| aCL | 45/71 | 29/205 | 32.420 | <0.001 |
| LA | 46/70 | 54/179 | 9.854 | 0.002 |

Abbreviations: TE: thromboembolism; ESR: erythrocyte sedimentation rate; CRP: C reactive protein; WBC, white blood cells; PLT: Platelet; APTT:activated partial thromboplastin time; FIB: fibrinogen; AT-III:Antithrombin III; TT:Thrombin time; FDP:Fibrin degradation products; ALP:Alkaline phosphatase; GGT:glutamyltranspeptidase; TB:Total bilirubin; TP:Total protein; AGR:Albumin globulin ratio; C3: complement C3; C4:complement C4; LA:lupus anticoagulant; aCL:Anti-cardiolipin; aβ2-GP I :Anti- β 2-glycoprotei I antibody.

### Multivariate Logistic Regression Analysis

Thromboembolism in ITP was used as the dependent variable for multivariate logistic regression analysis; variables that were significant (P < 0.1) in single factor analysis were used as concomitant variables. Multivariate logistic regression (the step-back method) showed that a high complement D2 level (odds ratio [OR]: 1.235; 95% confidence interval [CI]: 1.048–1.455), a high PLT level (OR: 1.015; 95% CI: 1.07–1.023) and a high Padua score (OR 1.190: 95% CI: 1.054–1.345) were identified as independent risk factors for thrombosis in ITP patients (Table 3).

**Table 3.** Multivariate Logistic Regression Analysis.

| Variable | B | S.E. | Wald | Sig. | Exp(B) | 95.0% C.I. for EXP(B) |  |
| --- | --- | --- | --- | --- | --- | --- | --- |
|  |  |  |  |  |  | Lower | Upper |
| Padua scores | .174 | .062 | 7.862 | .005 | 1.190 | 1.054 | 1.345 |
| D2 | .211 | .084 | 6.376 | .012 | 1.235 | 1.048 | 1.455 |
| PLT | .015 | .004 | 13.793 | .000 | 1.015 | 1.007 | 1.023 |
| Constant | -1.078 | .373 | 8.348 | .004 | .340 |  |  |

### Establishment and Evaluation of a Predictive Model

A predictive model for the risk of thromboembolism (Table 4) was established in line with the results arising from multivariate logistic regression analysis. The regression equation is as shown in Equation (1).Equation (1): Logit(P)= -1.078+ 0.015× complement PLE + 0.211× D2+0.174×Padua score. A ROC curve was generated from the model along with a separate Padua rating scale for predicting thrombus in AAV patients. DeLong inspection was used to compare the AUC of the model and the Padua rating scale(Figure 1). The AUC of the new risk prediction model was 0.673(95% CI: 0.615–0.730), thus, showing that the predictive performance of the model was good. The maximum Youden index, sensitivity and specificity were 0.272, 47.0% and 80.2%, The AUC of the Padua rating scale, PLT, D2, Padua scores was 0.637 (95% CI: 0.575–0.699),0.540 (95% CI: 0.478–0.602), 0.609 (95% CI: 0.548–0.669), respectively.(Table 4).

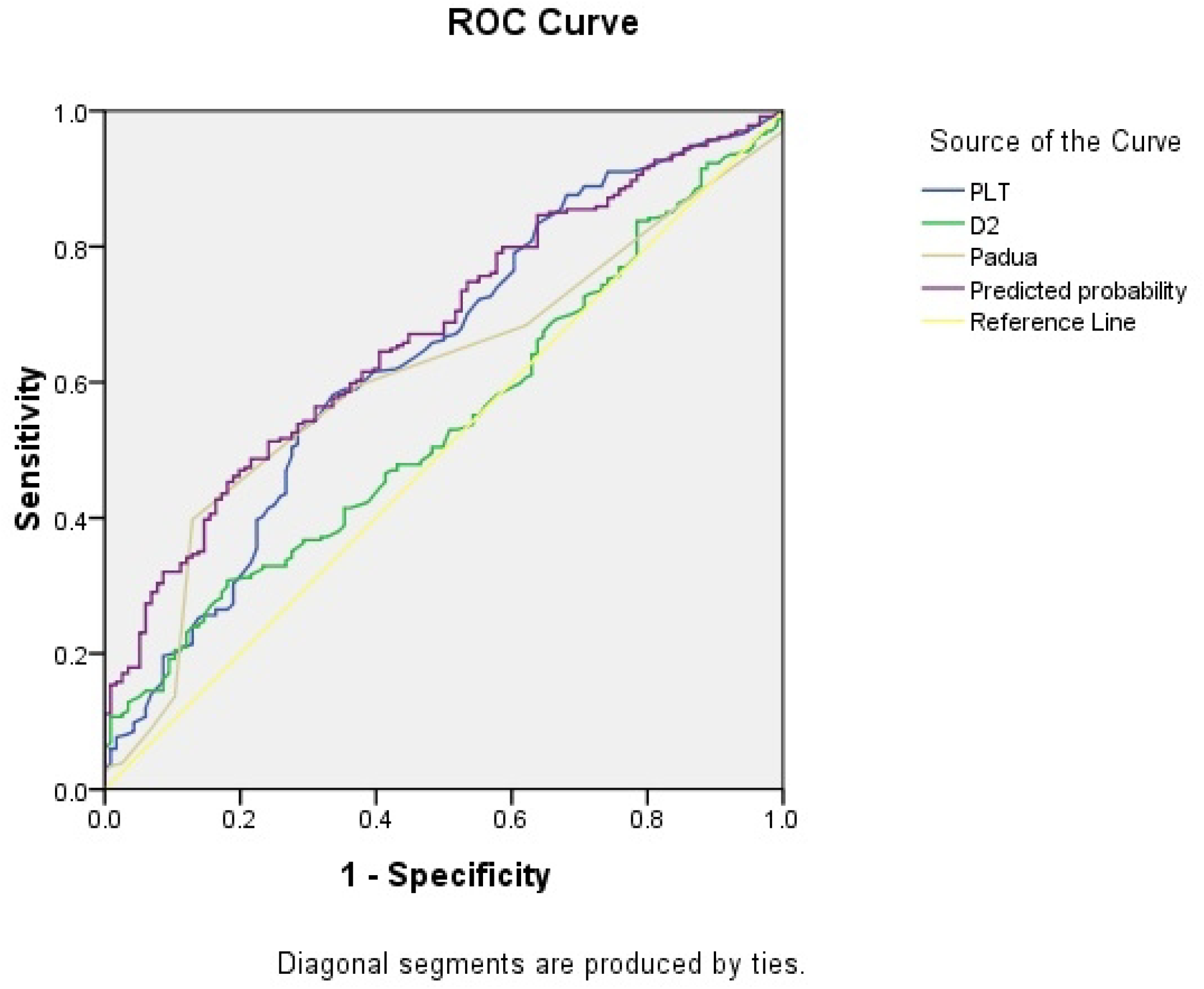

**Table 4.** Area Under the Curve.

| Variable | Area | S.E. | Sig. | Asymptotic 95% Confidence Interval |  |
| --- | --- | --- | --- | --- | --- |
|  |  |  |  | Lower Bound | Upper Bound |
| PLT | .637 | .032 | .000 | .575 | .699 |
| D2 | .540 | .032 | .228 | .478 | .602 |
| Padua scores | .609 | .031 | .001 | .548 | .669 |
| Predicted probability | .673 | .029 | .000 | .615 | .730 |

DeLong inspection showed that the new risk prediction model was significantly superior to the separate Padua rating scale (P = 0.033<0.05).(Table 5).

**Table 5.** DeLong-Pairwise sample region differences under the ROC curve.

| Variable | Z | Sig. | AUC | SE | 95% Confidence Interval |  |
| --- | --- | --- | --- | --- | --- | --- |
|  |  |  |  |  | Lower Bound | Upper Bound |
| Padua - PRE_1 | 2.135 | .033 | 0.064 | .243 | .005 | .123 |

## Discussion

Thrombosis risk factors are frequent in patients with ITP. Furthermore, the frequency of thrombosis risk factors is likely to be higher in patients with chronic ITP; long-term safety data from the fostamatinib trials showed that 58% of patients had multiple risk factors for thrombosis[13]. Therefore, given the prevalence of thrombosis risk factors in patients with ITP, it is important that risk factor assessment and risk minimization are continuously evaluated and managed throughout the course of the patient’s ITP history.Therefore, identifying potential risk factors for thrombosis will help us to develop a predictive model for the risk of thrombi so that we can identify high-risk patients in the early phases of disease and thus perform targeted interventions and prevent thrombosis.

Our results suggested that weight, ESR, Padua scores and D-dimer levels of patients in the ITP group with thrombi were significantly higher than those in the ITP group without thrombi. The levels of Neutrophil count, PLT count, FIB and FDP were significantly lower than those in the group without thrombi, indicating that these indicators might be potential risk factors for thromboembolic events. Thrombosis is associated with a reduced number of platelets. Although bleeding is one of the main symptoms of primary immune thrombocytopenia (ITP), risk factors for bleeding have yet to be fully established. Low platelet count PLT count; <20-30 × 10(9) /L) is generally indicative of increased risk of bleeding. However, PLT count and bleeding events cannot be fully correlated; many other patient- and disease-related factors are thought to contribute to increased bleeding risk. Furthermore, even though ITP patients have thrombocytopenia and are at increased risk of bleeding, ITP also carries higher risk of thrombotic events[14].

Our study has identified that severe thrombocytopenia and extremely high D-dimer can predict the risk of thrombosis. An increased level of D-dimer indicates that hypercoagulability and hyper-fibrinolysis have occurred and that the risk of thromboembolism has increased. In accordance with the present results, previous studies have demonstrated that anti-platelet factor 4 (PF4) antibodies, thrombocytopenia, high D-dimer, and hypofibrinogenemia can increase the risk of thrombosis for Vaccine-induced immune thrombocytopenia and thrombosis (VITT)[15].

Comorbidities, ITP treatments and aspects of ITP itself have been shown to play a role in this increased thrombosis risk. Evidence of dysregulated pro-/anti-inflammatory cytokines in ITP have suggested that ITP is an inflammatory disease and this increased inflammatory activity can lead to induced thrombosis. 34 Other factors of ITP itself with a potential prothrombotic role include a large proportion of young activated platelets and the presence of pro-coagulant, pro-inflammatory micro-particles.Both are elevated in patients with ITP, but definitive evidence linking them to increased thrombosis is lacking[16]. They highlighted that older age (>60),were independent predictors of both venous and arterial thrombotic events[17]. Therefore, it has been suggested that if we wish to prevent thrombosis in aged patients. However, our study has been unable to demonstrate that older age is a strong predictor for major thrombosis events. It may be that our study was not long enough, the follow-up was not long enough, and we will make improvements in the future.

Studies have indicated that 30% to 75% of ITP patients are accompanied by positive antiphospholipid antibodies [18]. Antiphospholipid antibodies encompass LA, aCL, and aβ2GP. Some studies also have suggested an association between aPLs and increased thrombotic risk[19]. The patients with positive aβ2-GPI, LA and aCL in the thrombus group was higher than that in the non-thrombotic group. Hence, it was concluded that the presence of aβ2-GPI antibodies, LA and anticardiolipin (aCL) increased the risk of thrombosis in ITP patients, especially the existence of LA. The mechanism by which positive antiphospholipid antibodies cause thrombosis may lie in the activation of platelet function after antibodies are coated with platelets. The antibody complex further activates the complement system (sensitization chemotactic effect), increasing the degree of platelet aggregation, which enhances the risk of thrombus formation[20]. At the same time, megakaryocytes can produce coagulation factors Ⅰ, Ⅷ, Ⅻ, and infantile megakaryocytes increase during the course of ITP pathogenesis, further inducing over-expression of coagulation factors, leading to hypercoagulability[21].

However, PC does not fully correlate with bleeding risk and there are multiple other risk factors thought to play a role[22]. Bleeding events can be difficult to predict, and some patients with a low PC, most of the time, display only mild bleeding symptoms[23].Even though ITP is a bleeding disorder, and despite the presence of thrombocytopenia, patients are also at a higher risk of venous and arterial thrombotic events. This risk of thromboembolism is increased in the presence of certain patient characteristics, such as older age, certain ITP treatments and even aspects of the disease itself[24]. Thrombotic events will also require initiation of anticoagulant and/or anti-platelet therapy and this can be challenging in patients with low PC.

A multi-factor analysis in this study showed that ITP patients with a high Padua score were more likely to develop thromboembolism. The Padua Predictive Score Scale is a thromboembolic risk assessment model first proposed by Professor Barbar in 2010[12]. The items of the Padua thrombosis scale are concise and clear; The scale is based on scientific projects and is widely used in clinical practice. The value of the Padova scale in predicting the risk of thrombosis has been confirmed by many studies. In this study, we found that a higher Padua score also had a certain predictive value for arteriovenous thrombosis in ITP patients, but further research is needed. Therefore, it is necessary to conduct dynamic Padua score measurement in ITP patients, and take corresponding preventive measures in time for patients with high score.

There are many factors for ITP patients to have thrombosis involving ITP itself, its treatment and the patients’ constitution, medical history, and former medication. ITP is not only a hemorrhagic disease but also a thrombotic disease. Clinicians should be alert to the risk of thrombotic diseases in ITP treatment. Therefore thrombus monitoring and screening should be carried out, and early prevention or appropriate anticoagulant treatment should be selected, especially for patients with high risk[25].

The receiver operating characteristic (ROC) curve is an important tool for the evaluation and comparison of predictive models when the outcome is binary. If the class membership of the outcomes is known, ROC can be constructed for a model, and the ROC with greater area under the curve indicates better performance[26]. ROC curve plots the true-positive rate (TPR) versus false-positive rate (FPR) under various thresholds for predictive results. Classifier with ROC having higher area under the curve (AUC) is usually more desirable. In our prediction model, the AUC was 0.673(95% CI: 0.615–0.730), the AUC of the Padua scores was 0.637 (95% CI: 0.575–0.699). Although the area of prediction model is 0.673 that is the largest in all variables, it is less than 0.7. Some studies have shown that the area greater than 0.5 indicates meaningful[27]. In future work, we plan to consider other more computationally efficient approaches for variance estimation, such as Taylor linearization variance estimation. Also, we plan on extending the estimation of the AUC for the ROC curves to estimation based on parametric and this will make model predictions more accurate.

Findings indicate that there could be an increased VTE risk with higher physical activity level including strenuous activities[28].Therefore, the exercise should be moderate and not excessive. For ITP patients, the nursing includes creating a comfortable hospitalization environment, maintaining appropriate temperature and humidity, and guiding family members to maintain quiet during visits. In addition, nursing staff need to regularly check the patient’s lower limbs, record skin color and temperature, inquire about discomfort such as swelling or pain, and promote VTE knowledge to the patient and their family. The patient’s diet is mainly light and high protein, encouraging them to eat more fresh fruits and vegetables to prevent constipation, and guiding them to exercise passively and actively. Specific measures include VTE risk assessment, psychological support, health education, disease monitoring, rehabilitation training, and intravenous care[29]. In summary, the nursing strategy offers a comprehensive treatment plan for patients with ITP. It effectively reduces the risk of VTE, improves coagulation function and blood routine indicators, enhances lower limb motor (LLM) function, and improves quality of life, gaining high patient recognition and helps prevent venous thromboembolism in these patients. Follow-up of patients after discharge is also very important, in terms of follow-up of the patient with a VTE diagnosis, it should be noted that, in the process of patient diagnosis/monitoring, immunology and hematology usually only takes care of the follow-up if patient has a ITP diagnosis. Through the follow-up, the nurse explains the precautions for the patient’s discharge and how to seek help if there is a problem, which will be more conducive to the recovery of the disease and improve the confidence in treatment.

Meanwhile, our study analyzed the risk factors of thromboembolism in ITP patients, constructed a thrombus risk prediction model and provided a method for the early identification of thrombosis in ITP patients to better help clinicians and nurse to actively prevent thrombosis in patients with ITP.

## Data Availability

All relevant data are within the manuscript and its Supporting Information files.

